# Polygenic prediction of cardiorespiratory fitness: The HUNT Study

**DOI:** 10.1101/2025.07.07.25330991

**Authors:** Karsten Øvretveit, Marie Klevjer, Ben M. Brumpton, Ulrik Wisløff, Kristian Hveem, Anja Bye

## Abstract

**Background:** Cardiorespiratory fitness (CRF) has a strong genetic component and low CRF is a major risk factor for cardiovascular morbidity and mortality. The purpose of this study was to develop and validate a polygenic score (PGS) for CRF (CRF_PGS_) and assess its associations with cardiovascular disease (CVD) and all-cause mortality. We hypothesized that the CRF_PGS_ would demonstrate similar cardioprotective benefits as the CRF phenotype.

**Methods:** Effect estimates from a genome-wide association study on directly measured CRF in the Trøndelag Health Study (HUNT; *n* = 4 525) were used in a Bayesian regression framework to develop multiple PGSs in an independent cohort from the UK Biobank (*n* = 65 165). The top performing score was applied in the HUNT target cohort, excluding the discovery sample (*n* = 82 109).

**Results:** The PGS-CRF association varied considerably as a function of model fit and phenotypic accuracy. In the target population, we observed a difference in CRF of 1.55 [95% confidence interval: 1.26, 1.84] mL·kg^-1^·min^-1^ between the bottom and top decile of the CRF_PGS_. Moreover, a high CRF_PGS_ demonstrated cardioprotective effects, with reduced risk for CVD, myocardial infarction, hypertension, and all-cause mortality. We also found that the CRF_PGS_ predisposed to lower risk of heart failure and hypertrophic cardiomyopathy in women.

**Conclusion:** A PGS for CRF derived from gold-standard phenotypes captures small, but potentially clinical meaningful variations in CRF, and is associated with reduced risk of cardiovascular morbidity and mortality. Heterogeneity in CRF phenotyping in large populations remains a challenge to PGS development and refinement.

## Introduction

Cardiorespiratory fitness (CRF), typically expressed as maximal (V^·^O_2max_) or peak (V^·^O_2peak_) oxygen uptake, has a strong and independent association with cardiovascular morbidity and mortality, with no apparent upper level of benefit (1–4). It is a complex phenotype that is influenced by multiple physiological parameters, including pulmonary, cardiovascular, and mitochondrial function (5). The performance of these intermediate phenotypes converges to set the individual’s upper limit of oxygen uptake.

The genetic component of V^·^O_2max_ is estimated to be between 44% and 68% from twin and family studies (6). V^·^O_2max_ is considered the gold standard measurement of CRF, and it is typically obtained by direct gas exchange analysis during a graded cardiopulmonary exercise test (CPET) (7). Although common practice in clinical research, CPET is less feasible in large cohort studies and often replaced by indirect measurements. The largest study with direct measurements of CRF to date was conducted as a part of the third wave of the Trøndelag Health Study (HUNT), where a total of 4 631 individuals completed a V^·^O_2peak_ test (8). Subsequently, a genome-wide association study (GWAS) in this population suggested 38 novel genetic variants associated with V^·^O_2peak_ (9). Functional annotation of the results confirmed previous genetic associations between variants predisposing to endurance, cardiac function, and cardiovascular disease (CVD).

A polygenic score aggregates genetic effect estimates from GWAS into a single metric that quantifies the liability to a specific trait. To date, no study has used gold standard measurements of CRF in polygenic prediction of the phenotype. Here, we use genetic and CRF data from multiple large independent cohorts to develop and test a PGS for CRF (CRF_PGS_). In addition to exploring the association between CRF_PGS_ and indices of CRF and related phenotypes, we assessed its impact on several CVD outcomes. We hypothesized that the CRF_PGS_ would be associated with estimated CRF (eCRF), and that it would demonstrate similar cardioprotective benefits as the CRF phenotype.

## Methods

### Participants

Beginning in 1984, HUNT has enrolled over 230 000 individuals in four separate surveys (10). Nearly 100 000 participants have contributed with biological material, of whom approximately 88 000 have provided DNA and consent for genotyping (11). Detailed information about genotyping is available in the supplemental material. We included up to 86 687 (45 970 women) genotyped and phenotyped participants from the HUNT study. The base data included individuals with directly measured V^·^O_2peak_ who had participated in the HUNT3 Fitness Study (*n* = 4 578, 2 343 women) (8, 9). To avoid overfitting, these were excluded from the HUNT target sample (*n* = 82 109, 43 627 women). The UK Biobank (UKB) is a prospective study of 500 000 participants from the United Kingdom that collects questionnaire data, clinical measurements, and biological samples. All participants in the UKB have been genotyped and imputed (12), and participants with genotype data, CRF, and physical activity (PA) measurements were included, resulting in a sample size of 65 165 (34 793 women).

### Ethics

The UKB research protocol and study design were approved by the NHS National Research Ethics Service, and all study participants provided written informed consent (project ID 40135). Ethical approval was obtained from the Northwest Centre for Research Ethics Committee (MREC, 11/NW/0382). In Scotland, the UKB has approval from the Community Health Index Advisory Group. The study was approved by the regional committee for medical research ethics (2019/29771), the Trøndelag Health Study, the Norwegian Data Inspectorate, and the National Directorate of Health. It was conducted in accordance with Norwegian laws and the Helsinki declaration. All participants provided written informed consent.

### Phenotyping

Disease outcomes were derived from health registry data spanning January 1999 through March 2020. Outcomes were defined in accordance with the 10th revision of the International Statistical Classification of Diseases and Related Health Problems (ICD-10) and grouped in disease categories (**Table S2**). In the HUNT3 Fitness Study, V^·^O_2peak_ was measured by trained personnel with a CPET (8). Self-reported outcomes were obtained from questionnaire data (**Table S3**). In HUNT4 participants, we also obtained body composition data from bioelectrical impedance analysis (InBody 770, Cerritos, CA, USA) and plasma hemoglobin (Hb). We calculated eCRF in HUNT3 and HUNT4 participants using a previously published nonexercise prediction model that incorporates age, waist circumference (WC), resting heart rate (RHR), and a PA-index (calculation available in the supplements) to determine sex-specific eCRF (13):

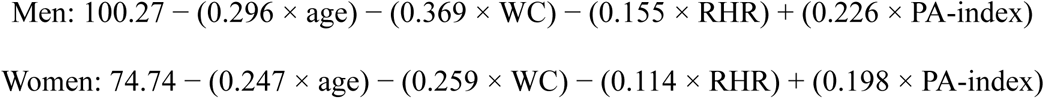

In the UKB, a submaximal bicycle test was used to assess CRF in a subsample of ∼100 000 participants. In short, the participants were divided into five groups by health risk, and the participants in the three healthiest groups were tested at 50%, 30% and at constant level of the predicted maximum workload. CRF was then determined as the net oxygen consumption based on body weight and maximum workload (14).

### Polygenic score

To develop a PGS for CRF, we used PRS-CS, a Bayesian regression framework with continuous shrinkage priors (15). The base data was the genotyped HUNT3 Fitness Study participants (9), and an independent UKB cohort was used as the tuning cohort. After testing multiple PGSs, the score with the strongest association with CRF was identified and subsequently applied in an independent HUNT target cohort (**Table S1**). A total of 1 104 501 SNPs (99.1%) overlapped between the PGS developed for CRF in the tuning and testing cohort (**Table 1**).

**Table 1.** Development flow of a polygenic score (PGS) for cardiorespiratory fitness (CRF)

| Type | Cohort | *n* SNPs | Purpose |
| --- | --- | --- | --- |
| Base data | HUNT3 Fitness study (*n* = 4 525) | 14 036 359 | Provide summary statistics of genetic effect estimates for CRF. |
| Tuning data | UK Biobank (*n* = 65 165) | 1 115 064* | Association testing of multiple PGSs with different tuning parameters. |
| Target data | HUNT1-4 (*n* = 82 109) | 1 104 501* | Validation and additional testing of the PGS on relevant outcomes. |

*HUNT, the Trøndelag Health Study; SNP, single-nucleotide polymorphism.*
*\* Restricted to HapMap3 variants.*

### Statistical analysis

Genotype-phenotype and phenotype-phenotype associations were assessed with the Pearson product-moment correlation coefficient. Disease risk estimates were derived from Cox proportional hazards models using the R package “survival” (16). We modelled sex-specific hazard ratios (HRs), adjusted for the first ten principal components of population structure, with age on the time scale. The odds of self-reported hyperglycemia and medication use were assessed with logistic regression models adjusted for age, age^2^, sex, and the first ten principal components. In both the survival and logistic models, the CRF_PGS_ was standardized to have a mean of “0” and standard deviation (SD) of “1” to estimate the effect of each SD change in the PGS. We also compared observed eCRF levels and mean change from HUNT3 to HUNT4 between the top and bottom deciles of the CRF_PGS_ distribution. Due to nonnormal data distributions, group differences were assessed with the Mann–Whitney U test. Results are presented with 95% confidence intervals (CI) and p-values where applicable. P-values were interpreted as continuous indicators of evidence strength and conclusions were drawn based on the magnitude and precision of effect sizes. Statistical analyses were performed with R (version 4.2.3).

## Results

### Participants characteristics and genotype-phenotype associations

The mean eCRF in HUNT3 and HUNT4 was 35.7 ± 8.2 mL·kg^-1^·min^-1^ and 35.0 ± 8.8 mL·kg^-1^·min^-1^, respectively. Genotype-phenotype correlations for CRF_PGS_ varied substantially between cohorts, with the strongest relationship being observed in participants who were part of the base data (**Table 2**). In this subset, the *r*^2^ between eCRF at HUNT3 and measured V^·^O_2peak_ was 0.68 (*n* = 3 474) and 0.58 for eCRF at HUNT4 (*n* = 3 026). The CRF_PGS_ also correlated with all eCRF predictors individually (**Table 3**). It was also inversely correlated with BMI in both HUNT3 (*r* = −0.062 [95% CI: −0.071, −0.052]) and HUNT4 (*r* = −0.060 [95% CI: −0.069, −0.051]). In HUNT4, we observed an inverse relationship between CRF_PGS_ and body fat mass (*r* = −0.076 [95% CI: −0.085, −0.067]) and visceral fat (*r* = −0.078 [95% CI: −0.087, −0.069]), but not fat-free mass.

**Table 2.** Genotype-phenotype associations for CRF_PGS_.

| Variable | Cohort | Sample size | Pearson's *r* | 95% CI |
| --- | --- | --- | --- | --- |
| Measured $\dot{V}O_{2peak}$ | HUNT3 Fitness study | 4 578 | 0.693 | 0.677, 0.707 |
| eCRF (HUNT3) | HUNT3 Fitness study | 3 474 | 0.345 | 0.315, 0.374 |
| eCRF (HUNT4) | HUNT3 Fitness study | 3 026 | 0.352 | 0.320, 0.383 |
| eCRF (HUNT3) | All HUNT | 32 381 | 0.113 | 0.102, 0.124 |
| eCRF (HUNT4) | All HUNT | 40 287 | 0.094 | 0.085, 0.104 |
| eCRF (HUNT3) | All HUNT w/o GWAS | 28 907 | 0.038 | 0.027, 0.050 |
| eCRF (HUNT4) | All HUNT w/o GWAS | 37 261 | 0.052 | 0.042, 0.062 |

*CI, confidence interval; HUNT, the Trøndelag Health Study; CRF<sub>PGS</sub>, polygenic score for cardiorespiratory fitness; $\dot{V}O_{2peak}$ , peak oxygen uptake; eCRF, estimated cardiorespiratory fitness; GWAS, genome-wide association study. Sample size reflect complete cases for both variables in each cohort.*

**Table 3.** Associations between CRF_PGS_ and eCRF predictors.

|  | HUNT3 |  | HUNT4 |  |
| --- | --- | --- | --- | --- |
|  | Pearson's *r* | 95% CI | Pearson's *r* | 95% CI |
| Age | 0.016 | 0.007, 0.025 | 0.013 | 0.004, 0.021 |
| Waist circumference | -0.054 | -0.063, -0.045 | -0.075 | -0.084, -0.066 |
| Resting heart rate | -0.035 | -0.045, -0.025 | -0.041 | -0.050, -0.032 |
| Exercise frequency | 0.043 | 0.034, 0.052 | 0.043 | 0.034, 0.052 |
| Exercise intensity | 0.021 | 0.010, 0.031 | 0.022 | 0.012, 0.032 |
| Exercise duration | 0.013 | 0.002, 0.023 | 0.020 | 0.010, 0.030 |
| Physical activity index | 0.030 | 0.019, 0.041 | 0.033 | 0.023, 0.043 |

*CRF<sub>PGS</sub>, polygenic score for cardiorespiratory fitness; eCRF, estimated cardiorespiratory fitness; HUNT, the Trøndelag Health Study; BMI, body mass index; CI, confidence interval. All coefficients derived from individuals who were not a part of the genetic base data.*

Correlation coefficients may be influenced by nonlinearities in the genotype-phenotype relationship, but the weighted average difference in eCRF between the bottom and top deciles of the CRF_PGS_ in HUNT3 (1.43 [95% CI: 1.01, 1.86] mL·kg^-1^·min^-1^, *n* = 2 891) and HUNT4 (1.66 [95% CI: 1.26, 2.06] mL·kg^-1^·min^-1^, *n* = 3 727) was 1.55 [95% CI: 1.26, 1.84] mL·kg^-1^·min^-1^ (**Figure 1**; **Table 4**).

**Figure 1.**
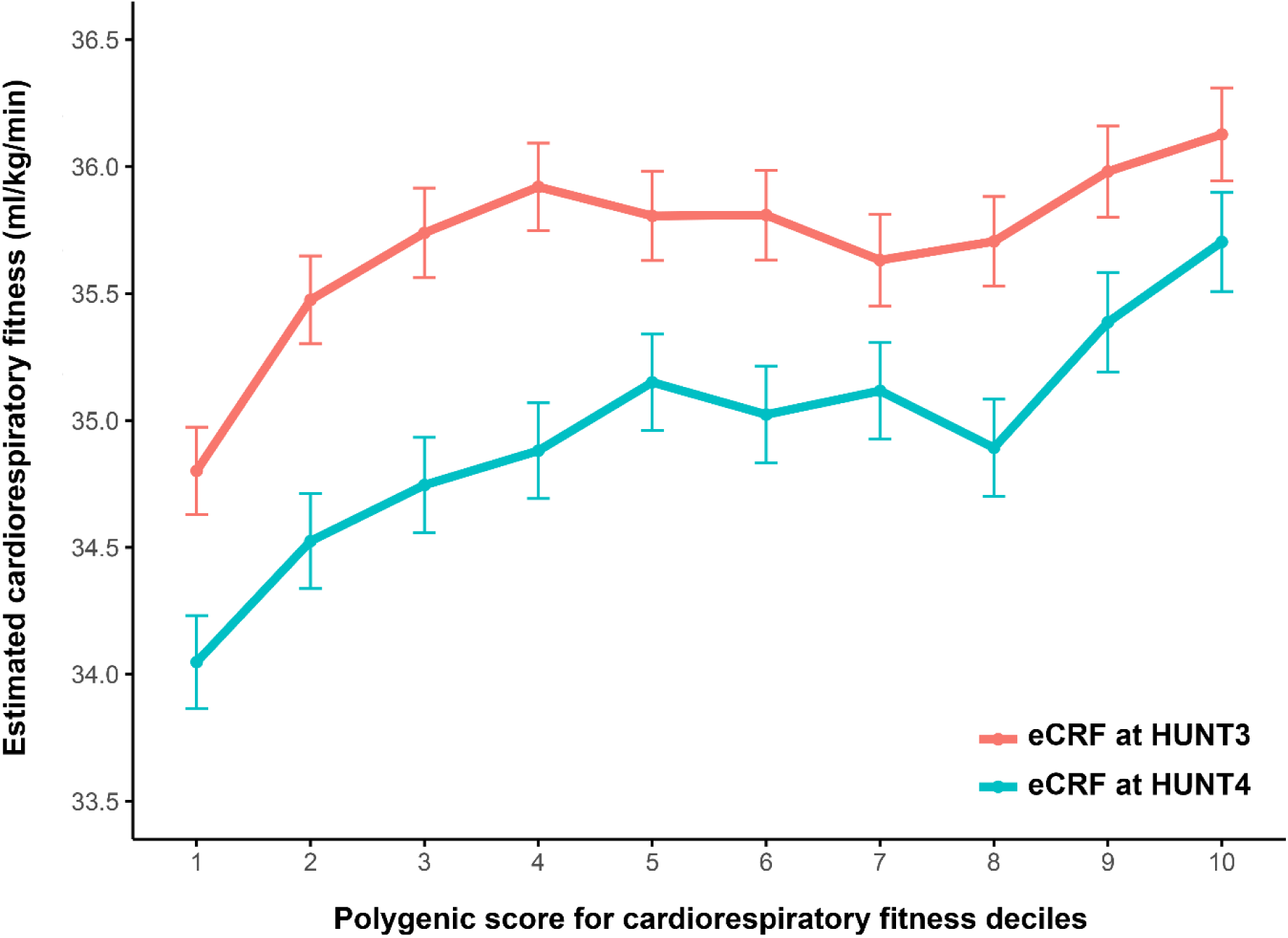
Cardiorespiratory fitness for each decile of the CRF_PGS_ at HUNT3 and HUNT4 *CRF_PGS_,* polygenic risk score for cardiorespiratory fitness; HUNT, the Trøndelag Health Study. Data presented as mean with 95% confidence intervals for each decile.

**Table 4.** Descriptive characteristics of the bottom and top decile of the CRF_PGS_ at HUNT3 and HUNT4.

|  | HUNT3 |  | HUNT4 |  |
| --- | --- | --- | --- | --- |
|  | Bottom decile | Top decile | Bottom decile | Top decile |
| Women/men | 1 687/1 204 | 1 618/1 273 | 2 185/1 542 | 2 063/1 663 |
| Age (years) | 53.9 ± 15.2 | 54.4 ± 16.4 | 50.9 ± 12.8 | 51.6 ± 13.4 |
| BMI (kg/m<sup>2</sup>) | 27.6 ± 4.5* | 26.4 ± 4.0* | 27.4 ± 4.7* | 26.4 ± 4.2* |
| eCRF (mL·kg<sup>-1</sup>·min<sup>-1</sup>) | 34.8 ± 8.0* | 36.2 ± 8.5* | 34.0 ± 8.5* | 35.7 ± 9.0* |
| SBP (mmHg) | 131 ± 19 | 131 ± 19 | 129 ± 18 | 128 ± 19 |
| DBP (mmHg) | 74 ± 11 | 73 ± 11 | 74 ± 10 | 73 ± 10 |
| PA index | 10.2 ± 9.2* | 11.1 ± 9.2* | 10.6 ± 9.4* | 11.6 ± 9.6* |

*HUNT, the Trøndelag Health Study; CRF<sub>PGS</sub>, polygenic score for cardiorespiratory fitness; BMI, body mass index; SBP, systolic blood pressure; DBP, diastolic blood pressure; PA index, physical activity index. Individuals with data on both CRF<sub>PGS</sub> and estimated cardiorespiratory fitness (eCRF) were included in the comparison; \* $p < 0.004$ (0.05/12) between deciles.*

Moreover, participants in the bottom decile of the CRF_PGS_ had a 7% greater drop in eCRF from HUNT3 to HUNT4 compared to those in the top decile (*p* = 0.04), indicating that the CRF_PGS_ may reflect preserved CRF throughout the lifespan. Although the CRF_PGS_ reflects phenotypic differences at the tails of the distribution, it appears to be less suited to identify individuals with the highest values (**Table 5**).

**Table 5.** Frequency of the top 100 eCRF values in each PGS decile.

| Decile | 1 | 2 | 3 | 4 | 5 | 6 | 7 | 8 | 9 | 10 |
| --- | --- | --- | --- | --- | --- | --- | --- | --- | --- | --- |
| *N* | 5 | 12 | 13 | 8 | 8 | 12 | 10 | 9 | 5 | 18 |
| Mean eCRF (mL·kg<sup>-1</sup>·min<sup>-1</sup>) | 62.2 | 64.1 | 63.0 | 63.4 | 63.2 | 63.7 | 64.2 | 63.0 | 63.4 | 63.0 |

*eCRF, estimated cardiorespiratory fitness; PGS, polygenic score.*

### PGS associations with disease outcomes

Survival models for multiple disease outcomes suggested that a genetically high CRF may offer cardiovascular protection (**Figure 2**). One SD increase in the CRF_PGS_ was associated with reduced incidence of CVD, myocardial infarction (MI), hypertension, and all-cause mortality for both sexes, as well as heart failure (HF) and hypertrophic cardiomyopathy (HCM) in women. Higher CRF_PGS_ was also associated with reduced frequency of antihypertensive and cholesterol-lowering medication, as well as self-reported hyperglycemia (**Figure 3**).

**Figure 2.**
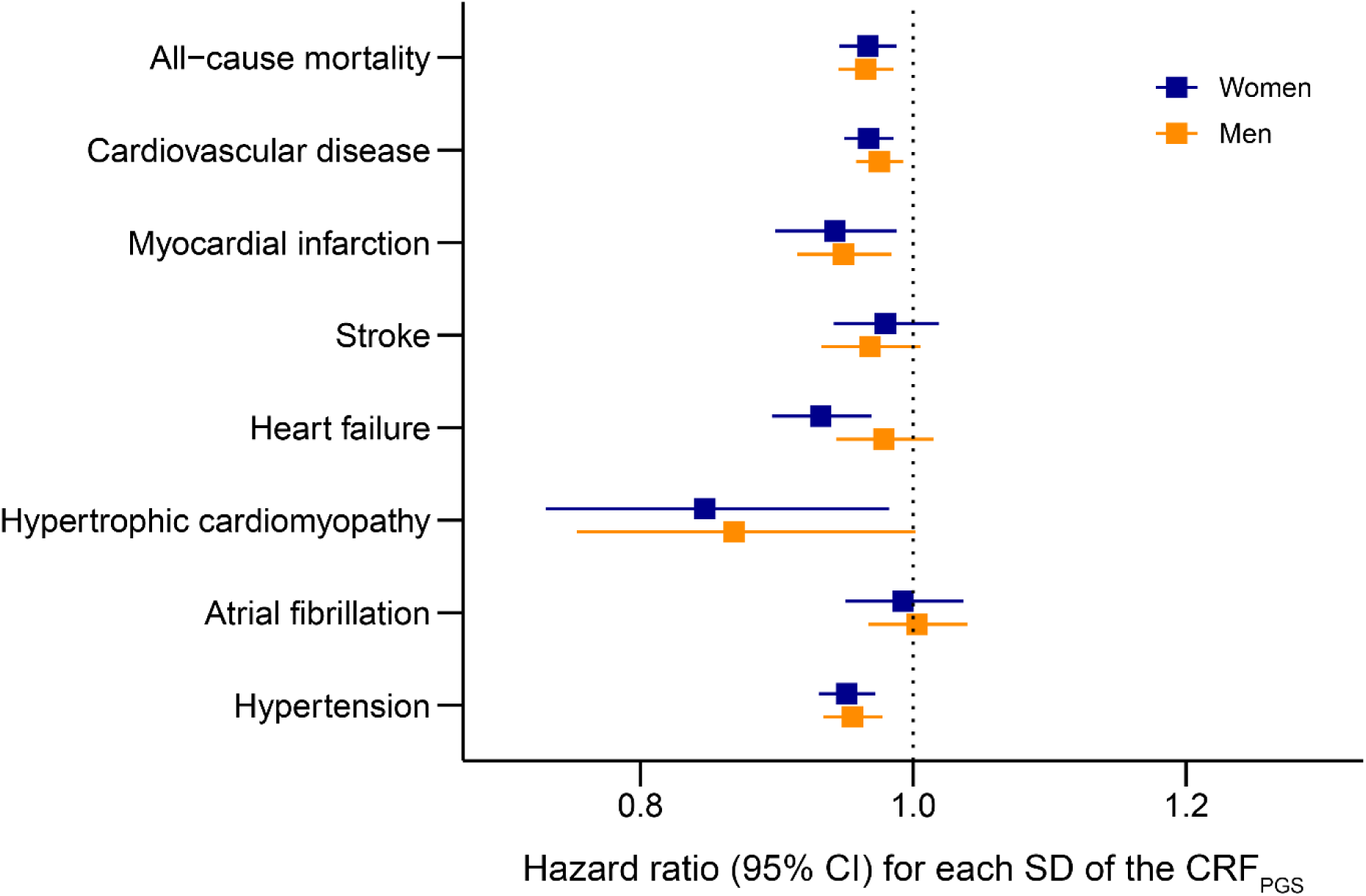
Cardiovascular disease risk CRF_PGS_, polygenic risk score for cardiorespiratory fitness; CI, confidence interval, SD; standard deviation. Disease outcomes were obtained from hospital records spanning January 1999 through March 2020 in participants from the Trøndelag Health Study that were not included in the base data (HUNT; n = 82 109). The models were adjusted for the first ten principal components with age as the time scale.

**Figure 3.**
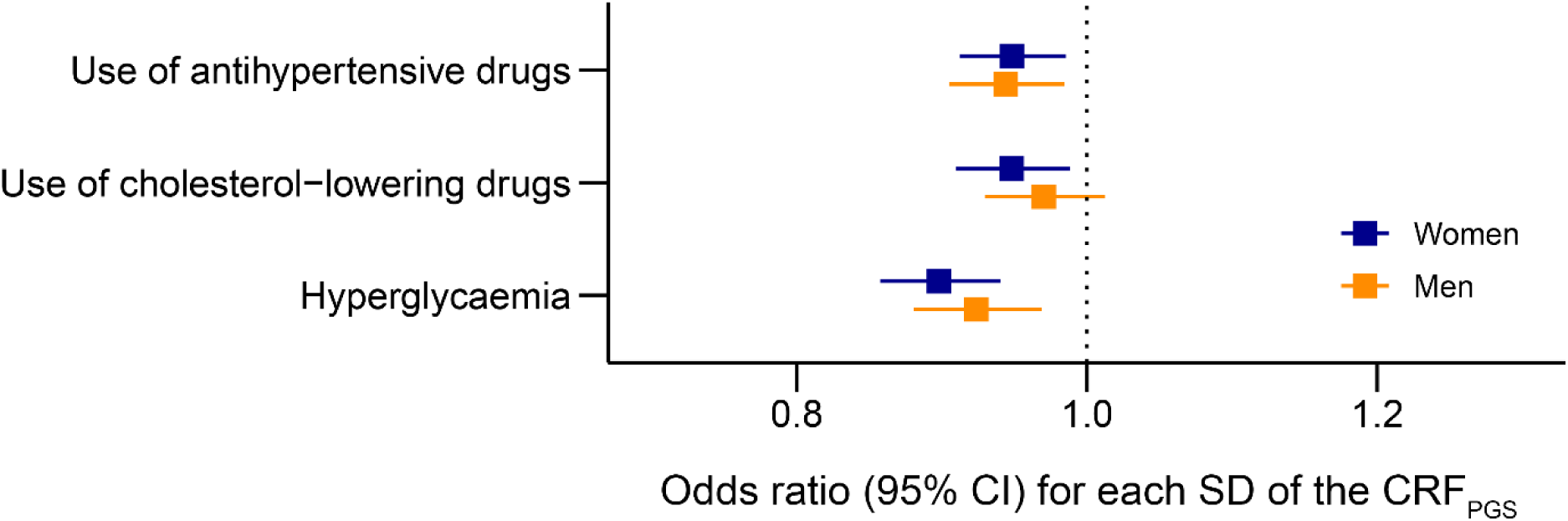
Odds of hyperglycemia and use of lipid-lowering and blood pressure drugs CRF_PGS_, polygenic risk score for cardiorespiratory fitness; CI, confidence interval; SD, standard deviation. The models were adjusted for age, age^2^, and the first ten principal components. The outcomes are derived from questionnaires distributed to participants in the latest Trøndelag Health Study (HUNT) survey (HUNT4) that were not included in the base data. The phrasing of each questionnaire item is available in the supplementary material.

We observed an association between eCRF and Hb at HUNT4 (*r* = 0.181 [95% CI: 0.171, 0.191]), but no such relationship was apparent between CRF_PGS_ and Hb (*p* = 0.11).

## Discussion

By using large, independent cohorts, we developed and validated the first genome-wide PGS for CRF using gold standard phenotypes as the base data. We found potentially clinically meaningful differences between high- and low-PGS carriers (17–19), as well as reduced risk of CVD for each SD increase in the CRF_PGS_. Although the phenotypic correlation was low in the target sample, our findings suggest that a genetic predisposition to elevated CRF is associated with reduced risk of all-cause mortality, CVD, MI, HF, HCM, and hypertension, as well as self-reported use of antihypertensive drugs, cholesterol drugs and hyperglycemia, suggesting shared genetics between CRF and these outcomes.

The pathogenesis of CVD is influenced by both genetic and environmental factors, and the genetic predisposition is highly heterogenic and complex (20, 21). Low CRF has previously been identified as an important risk factor for both all-cause mortality and CVD, and different subtypes of CVD including MI, hypertension, atrial fibrillation (AF), stroke, and HF (22). However, there are few studies investigating the genetic contribution to these relationships, mainly due to lack of large cohorts with directly measured CRF. The association between the CRF_PGS_ and ACM, CVD, MI, HF, HCM, and hypertension observed in the present study suggests shared genetics between CRF and these diseases. In a Swedish nationwide twin study they found indications that genetic factors influence the association of CRF with CVD and all-cause mortality, emphasizing the importance of the genetic contribution to CRF, independent of the protective effect of exercise-induced CRF (23).

The CRF_PGS_ was associated with lower risk of HF in women, but not in men, suggesting sex-specific differences in the overlapping genetic architecture between CRF and HF. The CRF_PGS_ was also associated with lower risk for HCM in women, albeit with some uncertainty around the estimate due to few events (372 cases). A similar, but weaker association was observed in men. Left ventricular function is one of the most important components of a high CRF(24). Furthermore, a reversal of left ventricular remodeling has been seen alongside exercise-induced improvements in V^·^O_2peak_ (25). The association between CRF_PGS_ and HCM risk may suggest that the CRF_PGS_ captures genetic signals related to cardiac function. Indeed, the genes *MYH7* and *MYBPC3* are responsible for nearly 70% of all HCM mutations, with the *MYH7* also being associated with CRF (26), indicating a genetic overlap with the CRF_PGS_. Patients with HCM are recommended to perform a CPET to assess the complex pathophysiology and severity of disease, as V^·^O_2peak_ has a great prognostic value for HCM patients (27). The CRF_PGS_ was not associated with either increased or decreased risk of AF. It has been shown that exercise has a U-shaped association with AF, meaning that both people with low and (extremely) high exercise levels are at increased risk (28, 29). Interestingly, a genetically high RHR appears to protect against AF despite increasing the risk of other cardiovascular diseases (30). Thus, although RHR is often used as a proxy for fitness status, the genetic contribution to RHR-associated AF risk is contradictory. The lack of association between the CRF_PGS_ and AF might indicate that the previously reported inverse relationship between CRF and AF risk could be due to exercise-induced improvements in CRF, rather than a genetically high CRF.

The CRF_PGS_ had a comparable and even slightly stronger associations with some of the individual predictors in the eCRF model than the eCRF itself, as well as anthropometric variables such as BMI and body fat mass. Thus, its cardioprotective benefits may in part be driven by a more favorable body composition. The lack of association between CRF_PGS_ and Hb could also indicate that it is not a direct predictor of oxygen transport capabilities, but rather a higher CRF as a function of lower body mass, in line with recent findings on the relationship between genetically predicted body composition and V^·^O_2max_ (31). Although the evidence on the relationship between genetically predicted V^·^O_2max_ and a more favorable anthropometric and metabolic profile is equivocal (31), our observation that a higher CRF_PGS_ is associated with lower BMI, body fat mass, and odds of hyperglycemia is consistent with the findings of others (26).

Because a PGS can be calculated at birth, it can serve as a proxy for lifetime exposure to beneficial or detrimental risk factors, e.g., high blood pressure or a high CRF. Although cross-sectional differences in phenotypic expression might be small, such as the ∼1.6 mL·kg^-1^·min^-1^ observed between the bottom and top decile of the CRF_PGS_ in the present study, this can meaningfully impact disease outcomes (17, 18). Notably, an increase of 1 mL·kg^-1^·min^-1^ in V^·^O_2peak_ is associated with a ∼15% decrease in mortality risk in men and women with coronary heart disease (19). The bottom decile of the CRF_PGS_ had a 7% greater drop in eCRF from HUNT3 to HUNT4 compared to the top decile, indicating that genetics may help preserve CRF throughout the lifespan and reduce disease risk. Indeed, small differences in cardiovascular traits between the tails of a polygenic risk distribution has been observed from the first years of life (32). If we consider the area under the CRF curve to be the main driver of CRF-mediated disease protection, a slightly elevated CRF due to genetics may offer considerable protection towards the later stages of life.

The top decile had higher PA indices and lower BMI, which indicates that the CRF_PGS_ also captures signals related to PA patterns and body composition. However, completely disentangling genetic and environmental effects in these risk constructs for phenotypes that are a product of both is challenging, and a PGS will typically also contain environmental effects on modifiable phenotypes (33).

Using independent cohorts for each step of PGS development is important to avoid overfitting (34). The lack of gold-standard CRF data in large cohorts makes the development of a CRF_PGS_ challenging. In our three populations, available phenotypes were directly measured V^·^O_2peak_ with CPET, estimated V^·^O_2peak_ from submaximal exercise data, and predicted V^·^O_2peak_ from nonexercise data. To provide the most accurate base data for the PGS calculation, we used effect estimates from a GWAS on directly measured participants previously published by our group (9). By performing association testing in all three cohorts, we illustrate how different the magnitude of genotype-phenotype correlation is. As expected, the CRF_PGS_ showed the best fit in the population the genetic effect estimates were derived from. It had the lowest phenotypic correlation in the UKB tuning cohort, but although the strength of the coefficient increased about 3-5 times in the HUNT target sample, the relationship remained weak. This was likely influenced by a nonlinearity in the genotype-phenotype relationship, as evident by the eCRF in the 5^th^ to 8^th^ decile of the CRF_PGS_. In addition, lack of validation cohorts with directly measured CRF and small sample size in the base data might have contributed to the low phenotypic correlation.

Strengths of the study include the use of independent cohorts to develop the CRF_PGS_, including the largest dataset on directly measured V^·^O_2peak_ in genotyped participants; the application of a novel Bayesian PGS framework to test multiple tuning parameters; and the use of registry-based disease outcomes with time to event. The study was limited by the intricate and highly polygenic genetic architecture of the CRF phenotype, which contributed to a low genotype-phenotype association in the target sample. Moreover, the reliance on estimated CRF, which only explains about ∼60% of trait variance for the nonexercise model and ∼50% (Pearson’s *r* range 0.68–0.74) for the submaximal exercise test (35), makes it challenging to validate and quantify the true predictive accuracy of the CRF_PGS_. Expressing V^·^O_2max_ relative to body weight (mL·kg^-1^·min^-1^) may also disproportionally penalize heavy individuals (36) and also explain the correlation between the CRF_PGS_, BMI, and indices of body fat. In addition to larger sample sizes with valid CRF phenotypes, elucidating the genetic basis of relevant intermediate phenotypes, e.g., left ventricular and mitochondrial function, may be required to more accurately predict CRF with genetic data.

## Conclusion

We developed and validated a genome-wide CRF_PGS_ using a Bayesian regression framework and large, genotyped cohorts. Although heterogeneity in CRF measurements across cohorts influenced the genotype-phenotype relationships, the CRF_PGS_ was associated with lower risk of CVD, MI, HF, HCM, hypertension, and all-cause mortality. Genetic susceptibility to higher CRF appears to lead to slightly elevated CRF within the normal range, rather than supraphysiological levels.

## Data Availability

Researchers affiliated to a Norwegian research institution can apply for HUNT data access from HUNT Research Centre (www.ntnu.edu/hunt) if they have obtained project approval from the Regional Committee for Medical and Health Research Ethics (REC). Researchers not affiliated to a Norwegian research institution should collaborate with and apply through a Norwegian principal investigator. Information on the application and conditions for data access is available at www.ntnu.edu/hunt/data. The HUNT Databank website provides a detailed overview of the available variables in HUNT (www.ntnu.edu/hunt/databank). Certain data from ancillary HUNT projects may be subject to a time-limited exclusivity provided to the researchers who have financed and conducted the data collection. Biologic material is available for analyses, information on procedures is found at the HUNT Biobank website (https://www.ntnu.edu/hunt/hunt-biobank). Data from the health registries are not kept by HUNT; instead, linkages between HUNT and registry data have to be made for each research project and require that the principal investigator has obtained project-specific approval for such linkage by REC and each registry owner.

## Acknowledgments

We are grateful to all participants in the Trøndelag Health Study (HUNT) and the UK Biobank (UKB) for their participation in this study. The HUNT Study is a collaboration between the HUNT Research Centre (Faculty of Medicine and Health Sciences, NTNU Norwegian University of Science and Technology), Trøndelag County Council, Central Norway Regional Health Authority, and the Norwegian Institute of Public Health. The HUNT genotype quality control and imputation has been conducted by the K.G. Jebsen Center for Genetic Epidemiology, Department of Public Health and Nursing, Faculty of Medicine and Health Sciences, NTNU. This research has been conducted using the UK Biobank Resource under Application Number 40135.

